# Dynamics of T cell responses to COVID-19 vaccines and breakthrough infection in people living with HIV receiving antiretroviral therapy

**DOI:** 10.1101/2024.03.08.24304006

**Authors:** Sneha Datwani, Rebecca Kalikawe, Rachel Waterworth, Francis M. Mwimanzi, Richard Liang, Yurou Sang, Hope R. Lapointe, Peter K. Cheung, F. Harrison Omondi, Maggie C. Duncan, Evan Barad, Sarah Speckmaier, Nadia Moran-Garcia, Mari L. DeMarco, Malcolm Hedgcock, Cecilia T. Costiniuk, Mark Hull, Marianne Harris, Marc G. Romney, Julio S.G. Montaner, Zabrina L. Brumme, Mark A. Brockman

## Abstract

**Introduction:** People living with HIV (PLWH) can exhibit impaired immune responses to vaccines. Accumulating evidence indicates that PLWH, particularly those receiving antiretroviral therapy, mount strong antibody responses to COVID-19 vaccination, but fewer studies have examined cellular immune responses to vaccination. We measured SARS-CoV-2 spike-specific CD4+ and CD8+ T cell responses generated by two and three doses of COVID-19 vaccine in PLWH receiving antiretroviral therapy, compared to control participants without HIV. We also quantified T cell responses after post-vaccine breakthrough infection, and receipt of fourth vaccine doses, in a subset of PLWH.

**Methods:** We quantified CD4+ and CD8+ T cells reactive to overlapping peptides spanning the ancestral SARS-CoV-2 spike protein in 50 PLWH and 87 controls without HIV, using an activation induced marker (AIM) assay. All participants remained SARS-CoV-2 naïve until at least one month after their third vaccine dose. SARS-CoV-2 infection was determined by seroconversion to nucleocapsid (N) antigen, which occurred in 21 PLWH and 38 controls post-third dose. Multivariable regression analyses were used to investigate relationships between sociodemographic, health and vaccine-related variables and vaccine-induced T cell responses, as well as breakthrough infection risk.

**Results:** A third vaccine dose boosted spike-specific CD4+ and CD8+ T cell frequencies significantly above those measured after the second dose (all p<0.0001). Median T cell frequencies did not differ between PLWH and controls after the second dose (p>0.1), but CD8+ T cell responses were modestly lower in PLWH after the third dose (p=0.02), an observation that remained significant after adjustment for sociodemographic, health and vaccine-related variables (p=0.045). In PLWH who experienced breakthrough infection, median T cell frequencies increased even higher than those observed after three vaccine doses (p<0.03), and CD8+ T cell responses in this group remained higher even after a fourth vaccine dose (p=0.03). In multivariable analysis, the only factor associated with increased breakthrough infection risk was younger age, consistent with the rapid increases in SARS-CoV-2 seropositivity among younger adults in Canada after the initial appearance of the Omicron variant.

**Conclusion:** PLWH receiving antiretroviral therapy mount strong T cell responses to COVID-19 vaccines that can be enhanced by booster doses or breakthrough infection.

## INTRODUCTION

Prior to the widespread availability of COVID-19 vaccines, there was evidence that people living with HIV (PLWH) were at increased risk of severe COVID-19 outcomes [1–5], making vaccination particularly important in this group. Since then, a growing body of evidence indicates that COVID-19 vaccines have been highly effective at preventing severe disease associated with SARS-CoV-2 infection, including in PLWH [6, 7]. Studies analyzing vaccine-induced immune responses, including from our group, also confirm that PLWH, in particular those who are receiving suppressive antiretroviral therapy, mount robust humoral (antibody) response to COVID-19 immunization [8–16]. But comparably fewer studies have investigated the cellular immune response to COVID-19 vaccines in PLWH, which includes CD4+ helper T cells that play a central role in generating antigen-specific B cells and antibodies and CD8+ cytotoxic T cells that recognize and eliminate virus-infected cells [17–20]. Indeed, while COVID-19 vaccines typically induce strong T cell responses [21, 22], the frequency of spike-specific CD4+ T cells following COVID-19 vaccination may be lower in PLWH, in particularly among those with low CD4+ T cell counts [9, 23]. A better understanding of T cell responses elicited by COVID-19 vaccines, as well as breakthrough SARS-CoV-2 infections, will inform ongoing efforts to enhance protective immunity in PLWH.

Towards this goal, we investigated the dynamics of spike-specific CD4+ and CD8+ T cell responses elicited after two and three doses of COVID-19 vaccine in a cohort of 50 adult PLWH receiving antiretroviral therapy, and 87 controls without HIV. All participants remained naïve to SARS-CoV-2 until at least one month after their third dose. We additionally investigated spike-specific CD4+ and CD8+ T cell responses in a subset of 21 PLWH who experienced their first SARS-CoV-2 breakthrough infection between one and six months after their third vaccine dose. Overall, our results revealed that spike-specific CD4+ and CD8+ T cell frequencies were boosted with each vaccine dose, and that these responses were broadly comparable between PLWH and controls, with the exception of CD8+ T cell responses after the third dose, which were moderately lower in PLWH compared to controls. Breakthrough infection further boosted median spike-specific T cell frequencies, and CD8+ T cell responses remained higher in these individuals even after a fourth vaccine dose.

## METHODS

### Participants

Our cohort of COVID-19 vaccine recipients, based in British Columbia (BC) Canada, has been described previously [10, 13, 24]. Of the 99 PLWH and 152 controls (without HIV) originally enrolled in the study, 62 (PLWH) and 117 (controls) remained naïve for SARS-CoV-2 infection until at least one month after their third COVID-19 vaccine dose. Of these, we examined a subset of 50 PLWH and 87 controls, selected largely based on sample availability (**Table 1**).

**Table 1:** Participant characteristics.

| Characteristic | PLWH<br>(n=50) | Controls<br>(n=87) |
| --- | --- | --- |
| **Sociodemographic and health variables<sup>a</sup>** |  |  |
| Age in years, median [IQR] | 58 [42-65] | 50 [35-72] |
| Female sex at birth, n (%) | 6 (12%) | 58 (67%) |
| Non-white ethnicity, n (%) | 14 (28%) | 37 (43%) |
| Number of chronic health conditions, median [IQR]<sup>b</sup> | 1 [0-1] | 0 [0-1] |
| HIV plasma viral load (RNA copies per mL) | <50 [<50-<50] | N.A. |
| Recent CD4 cell count (per mm<sup>3</sup>), median [IQR] | 695 [468-983] | N.A. |
| Nadir CD4 cell count (per mm<sup>3</sup>), median [IQR] | 250 [140-490] | N.A. |
| **Vaccine details** |  |  |
| Initial regimen<sup>c</sup> |  |  |
| mRNA only, n (%) | 42 (84%) | 84 (97%) |
| ChAdOx1-containing, n (%) | 8 (16%) | 3 (3%) |
| Third dose |  |  |
| BNT162b2, n (%) | 17 (34%) | 34 (39%) |
| mRNA-1273, n (%) | 33 (66%) | 53 (61%) |
| Days between first and second doses, median [IQR] | 59 [53-68] | 89 [73-98] |
| Days between second and third doses, median [IQR] | 182 [141-191] | 197 [169-215] |
| **Post-3rd vaccine dose SARS-CoV-2 infections, n (%)<sup>d</sup>** | **21 (42%)** | **38 (44%)** |

<sup>a</sup>Sociodemographic, health, and vaccine data were collected by self-report and confirmed through medical records where available. CD4+ T cell counts and HIV plasma viral loads were obtained from medical records.
<sup>b</sup>Chronic conditions were defined as hypertension, diabetes, asthma, obesity, chronic diseases of lung, liver, kidney, heart or blood, cancer, and immunosuppression due to chronic conditions or medication.
<sup>c</sup>Initial regimen refers to the first two doses. "mRNA only" refers to participants who received BNT162b2 and/or mRNA-1273 while "ChAdOx1-containing" refers to participants who received two ChAdOx1 doses, or one ChAdOx1 dose plus one mRNA dose.
<sup>d</sup>SARS-CoV-2 infections that occurred between one and six months after the third vaccine dose.

### Ethics approval

Written informed consent was obtained from all participants. This study was approved by the University of British Columbia/Providence Health Care and Simon Fraser University Research Ethics Boards.

### SARS-CoV-2 seroconversion

SARS-CoV-2 infection was identified based on the development of serum antibodies against the viral Nucleocapsid (N) protein using the Elecsys Anti-SARS-CoV-2 assay (Roche Diagnostics), combined with diagnostic (PCR- and/or rapid-antigen-test) information where available.

### T cell activation induced marker assays

Cryopreserved peripheral blood mononuclear cells (PBMC) were thawed and diluted in TexMACS media (Miltenyi Biotec, Cat#130-097-196). PBMC were stimulated at 1 x 10^6^ cells per well for 24 hours with peptide pools spanning the SARS-CoV-2 ancestral spike protein (15-mers, overlapping by 11 amino acids) (Miltenyi Biotec, Cat#130-127-953) in duplicate in a 96-well U-bottom plate.

PBMC were incubated with media only (no peptide) as a negative control or 2 µg/ml CytoStim reagent (Miltenyi Biotec, Cat#130-092-172) as a positive control. Following stimulation, cells were labeled with CD8-APC/Cyanine7 (Biolegend, Cat#301016), CD4-FITC (Biolegend, Cat#300538), CD137-APC (Biolegend, Cat#309810), CD69-PE (BD, Cat#555531), OX40-PE-Cy7 (Biolegend, Cat#350012), CD3-PerCP/Cyanine5.5 (Biolegend, Cat#317336) CD14-V500 (BD, Cat#561391), CD19-V500 (BD, Cat#561121) and 7-AAD Viability Staining Solution (Biolegend, Cat#420404). Data were acquired on a Beckman Coulter Cytoflex S flow cytometer, with a minimum of 10,000 CD3+ T cells assayed per participant. After identifying CD3+CD4+ and CD3+CD8+ T cell subsets, the percentage of stimulated cells was determined based on upregulation of two activation markers, using CD137 and OX40 for CD4+ T cells, and CD137 and CD69 for CD8+ T cells (see gating strategy in **Figure S1**). Data were analyzed in FlowJo version 10.8.1.

### SARS-CoV-2 neutralization

We previously reported the ability of participant plasma to neutralize ancestral SARS-CoV-2 (isolate USA-WA1/2020; BEI Resources, Cat #NR-52281), based on the results of a live-virus assay [24].

### Statistical analyses

Continuous variables were compared using the Mann-Whitney U-test (for unpaired data) or Wilcoxon test (for paired measures). Relationships between continuous variables were assessed using Spearman’s correlation. Zero-inflated beta regressions were used to investigate the relationship between sociodemographic factors and vaccine-induced T cell responses using a confounder model that adjusted for variables that could influence these responses, or that differed in prevalence between groups. These regressions model the response variable as a beta-distributed random variable whose mean is given by a linear combination of the predictor variables (after a logit transformation). Beta distributions are bounded below and above by 0 and 100%, making this a standard choice of regression for frequency data. Because a beta distribution does not admit values of 0 (or 100), we used a zero-inflated beta distribution, which allows for zeros in the data, to accommodate the minority of non-responders (*i.e.* 0 values) in our data. For analyses performed after two-dose vaccination, included variables were: HIV status (non-PLWH as reference), age (per year), sex at birth (female as reference), ethnicity (non-white as reference), chronic conditions (per condition), ChAdOx1-containing initial vaccine regimen (mRNA only vaccination as reference) and the interval between first and second doses (per day). Analyses performed after three-dose vaccination also included the third COVID-19 mRNA dose brand (BNT162b2 as reference), the interval between second and third doses (per day), and the % spike-specific T cells after two doses (per percent increase). Multivariable logistic regression was used to explore the relationship between these same variables and the risk of SARS-CoV-2 breakthrough infection between one and six months after the third vaccine dose. All tests were two-tailed, with p<0.05 considered statistically significant. Analyses were conducted using Prism v9.2.0 (GraphPad) and in R.

## RESULTS

### Participant characteristics

Characteristics of the 50 PLWH and 87 controls without HIV are shown in **Table 1**. All participants remained COVID-19 naïve until at least one month after their third vaccine dose, as confirmed by repeated negative serology for SARS-CoV-2 anti-N antibodies, and lack of reporting of any SARS-CoV-2 positive test result (either by PCR or rapid antigen test), up to this time point. PLWH and controls were a median age of 58 (interquartile range [IQR] 42-65) and 50 (IQR 35-72) years old, respectively, and predominantly of white ethnicity (72% of PLWH and 57% of controls). Most PLWH were male (88%, reflecting the overall population of PLWH in BC), while most controls were female (67%, broadly reflecting the overall demographics of health care workers in the region, from which the controls were predominantly recruited). Both groups reported few chronic health conditions (excluding HIV infection; median 1 [IQR 0-1] in PLWH, versus median 0 [IQR 0-1] in controls). All PLWH were receiving combination antiretroviral therapy, with a median HIV plasma viral load of <50 copies/mL (IQR <50 - <50 copies/mL) at study entry. The median CD4+ T cell count at study entry among PLWH was 695 (IQR 468-983) cells per mm^3^, while median nadir CD4+ T cell count was 250 (IQR 140-490) cells per mm^3^.

Most participants (84% of PLWH and 97% of controls) initially received two doses of mRNA vaccine, either BNT162b2 or mRNA-1273; the remainder received at least one ChAdOx1 dose in their initial series. In PLWH, second vaccine doses were administered a median of 59 days after the first, compared to an median interval of 89 days in controls (p<0.0001). This is because Canada delayed second doses well beyond the manufacturers’ recommended 21-28 day interval, due to initially limited availability of vaccine supplies in the country [25]. As a result, individuals who received their first doses early in the vaccination campaign, including most health care workers, generally waited the longest to receive their second doses. Retrospectively, it was determined that delaying second doses did not reduce vaccine efficacy, and in some cases may have enhanced protection [26] and reduced the risk of myocarditis [27], prompting the World Health Organization to recommend a 2-month (∼60-day) interval between first and second doses [28]. As a result, our multivariable analyses adjust for between-dose intervals and other sociodemographic and clinical attributes listed in **Table 1**. Third vaccine doses were predominantly mRNA-1273 (66% of PLWH and 61% of controls) and were administered a median of 182 to 197 days (approximately 6 to 6.5 months) after the second dose. Third mRNA-1273 dose amounts also differed by age and health history: according to vaccine guidelines in British Columbia, adults 65 years of age and older, as well as PLWH with a history of immunodeficiency, were eligible for a 100 mcg dose whereas all others were eligible for a 50 mcg booster dose (all BNT162b2 doses were 30 mcg). A total of 21 (42%) of PLWH and 38 (44%) of controls experienced their first SARS-CoV-2 infection between one and six months after the third vaccine dose, as determined by seroconversion to the SARS-CoV-2 N protein and/or self-reported positive SARS-CoV-2 test results where available. Though viral genotyping was not performed for these “breakthrough” infections, these were likely Omicron BA.1 or BA.2 based on local molecular epidemiology trends at the time [29].

### T cell responses following two and three COVID-19 vaccine doses

Prior to vaccination, the percentage of spike-specific CD4+ and CD8+ T cells, measured in a subset of 10 PLWH and 15 controls, was negligible as expected (**Figures 1A, 1B**). Following two vaccine doses, the percentage of spike-specific CD4+ T cells increased significantly from pre-vaccine levels (paired measures p≤0.004 for both PLWH and controls), reaching a median 0.25% (IQR 0.07-0.50) in PLWH and a median 0.32% (IQR 0.15-0.55) in controls, a difference that was not statistically significant between groups (p=0.11; **Figure 1A**). CD8+ T cell responses also increased significantly from pre-vaccine levels following two vaccine doses (paired measures p≤0.002 in both groups), reaching a median 0.39% (IQR 0.22-0.53) in PLWH and a median 0.31% (IQR 0.14-0.61) in controls, a difference that was not statistically significant between groups (p=0.39; **Figure 1B**). The lack of statistically significant differences in vaccine-induced T cell responses between PLWH and controls remained after controlling for socio-demographic, health and vaccine-related variables (p=0.08 for CD4+ T cell responses; p=0.99 for CD8+ T cell responses; **Tables S1 and S2**). Of note, these multivariable analyses did not identify any variables that were significantly associated with spike-specific CD4+ T cell responses after two vaccine doses (**Table S1**), though having received at least one ChAdOx1 dose as part of the initial vaccine series was weakly associated with higher spike-specific CD8+ T cell responses after the second vaccine dose (p=0.049; **Table S2**).

**Figure 1.**
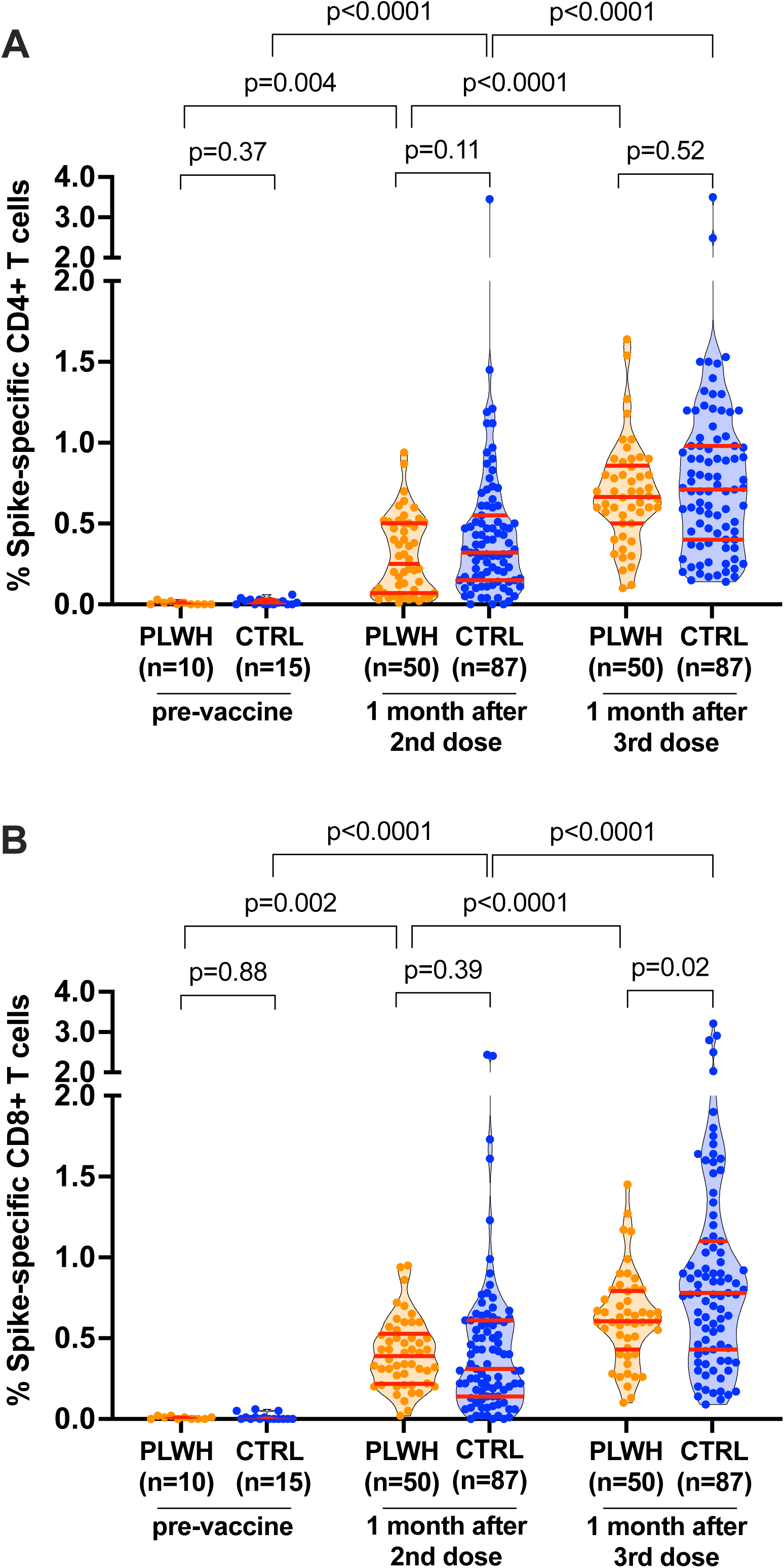
SARS-CoV-2 spike-specific T cell responses after COVID-19 vaccination. *Panel A:* Spike-specific CD4+ T cell frequencies before and after two and three-dose COVID-19 vaccination. People living with HIV (PLWH) are in orange; controls without HIV (CTRL) are in blue. All participants are COVID-19 naïve. Red bars indicate median and IQR. The Mann-Whitney U-test was used for between-group comparisons and the Wilcoxon matched pairs test was used for longitudinal paired comparisons. P-values are not corrected for multiple comparisons. *Panel B:* Same as panel A, but for spike-specific CD8+ T cell frequencies.

A third vaccine dose further boosted the percentage of spike-specific CD4+ T cells beyond the levels observed after two doses (paired measures p<0.0001 for both PLWH and controls; **Figure 1A**). After the third dose, spike-specific CD4+ T cell frequencies reached a median 0.67% (IQR 0.50-0.86) in PLWH compared to a median 0.71% (IQR 0.40-0.98) in controls (p=0.52; **Figure 1A**), where this lack of significant difference between groups remained after controlling for sociodemographic, health and vaccine-related variables (p=0.58; **Table S1**). Similarly, a third vaccine dose further boosted the percentage of spike-specific CD8+ T cells (paired measures p<0.0001 for both groups; **Figure 1B**). Somewhat in contrast to the other immune measures however, the median CD8+ T cell response in PLWH (0.61%; IQR 0.43-0.79) was lower compared to that of controls (0.78%; IQR 0.43-1.1) after the third vaccine dose (p=0.02; **Figure 1B**). This difference remained marginally statistically significant after controlling for sociodemographic, health and vaccine-related variables (p=0.045; **Table S2**).

The multivariable analyses also revealed that by far the strongest predictors of spike-specific CD4+ and CD8+ T cell responses after three vaccine doses were the corresponding percentage of spike-specific T cells achieved after two doses (**Tables S1, S2**). For example, the zero-inflated beta regression estimates for the impact of each 1% increment in post-second dose T cell frequencies on post-third dose frequencies were 0.71 for CD4+ T cells (p<2×10^-16^) and 0.79 for CD8+ T cells (p<2×10^-16^). To interpret these estimates: if the value of the linear predictor was x for a given set of values of the predictors, and the % of spike-specific CD4+ T cells after two vaccine doses was increased by 1%, the resulting value of the linear predictor would now be x + 0.71, which translates to a non-linear increase in the regressed mean from 100*logit(x) to 100*logit(x + 0.71), where the factor of 100 converts the proportions to percentages. Male sex, receipt of an mRNA-1273 booster, and, somewhat unexpectedly, a higher number of chronic health conditions, were also independently associated with higher spike-specific CD4+ T cell responses (estimates 0.11 - 0.24; all p<0.04; **Table S1**). We hypothesize that the association between chronic health conditions and better T cell responses is because, after adjusting for post-second-dose responses, individuals with such conditions benefited particularly from a third dose. The association with mRNA-1273 is likely because adults aged 65 years and older, as well as PLWH with a history of immune dysfunction were eligible for the higher (100mcg) mRNA-1273 third dose in British Columbia, whereas the general population received the standard 50 mcg booster dose.

### Correlations between vaccine-induced T cell responses and other immune measures

The percentages of spike-specific CD4+ and CD8+ T cells were strongly associated with one another after the second vaccine dose (Spearman’s ρ=0.57; p<0.0001) as well as after the third dose (Spearman’s ρ=0.54; p<0.0001; **Figure 2A**). No association was observed between the percentage of spike-specific CD4+ T cells and the magnitude of vaccine-induced neutralizing antibody responses based on a live-virus assay (reported previously by our group [24]) after either the second (Spearman’s ρ=-0.04, p=0.7) or third dose (Spearman’s ρ=-0.07, p=0.4; **Figure 2B**). Prior studies have linked the frequency of CD4+ T follicular helper cells with antibody responses [30], but we did not assess T cell subsets here.

**Figure 2.**
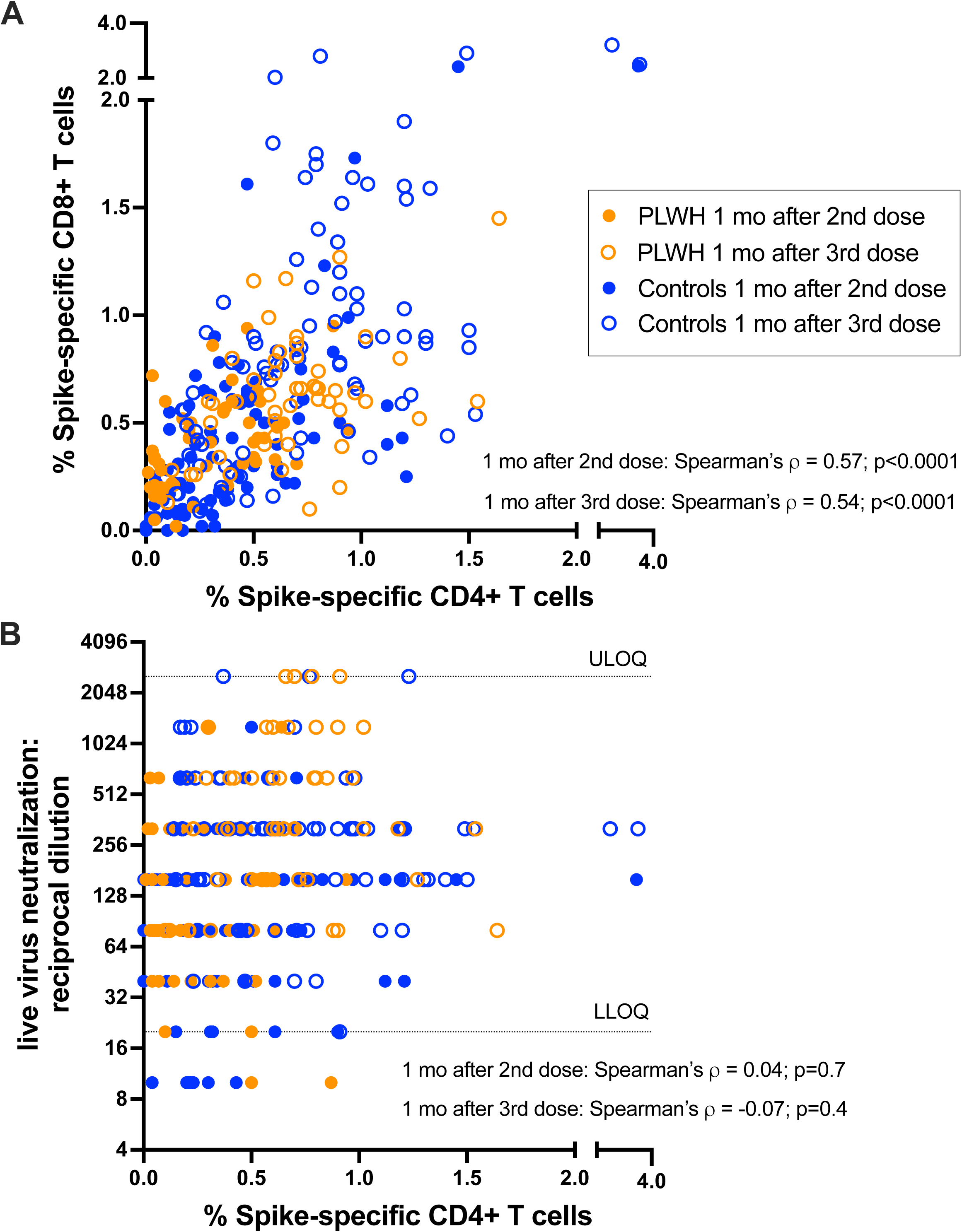
Associations between cellular and humoral immune measures after COVID-19 vaccination. *Panel A:* Correlation between spike-specific CD4+ and CD8+ T cell frequencies one month after the second dose (open circles) and one month after the third dose (closed circles) in people living with HIV (PLWH, in orange) and controls without HIV (in blue). *Panel B*: Correlation between spike-specific CD4+ T cell frequencies and SARS-CoV-2 neutralizing antibodies, measured at one month after the second dose (open circles) and one month after the third dose (closed circles) in people living with HIV (PLWH, in orange) and controls without HIV (in blue). All participants are COVID-19 naïve.

As CD4+ T cell counts are used as an indicator of immune function in PLWH, we evaluated the relationship between this parameter and vaccine-induced T cell responses in this group. We observed no association between the most recent CD4+ T cell count and the frequency of either spike-specific CD4+ or CD8+ T cells following the second or third vaccine doses (Spearman’s ρ=-0.09 to −0.18; all p>0.2; **Figure S2**). Similarly, we observed no association between nadir CD4+ T cell counts and vaccine-induced CD4+ T cell responses after either dose (Spearman’s ρ=-0.05 and −0.12, respectively, both p>0.4) (**Figure S2**). By contrast, an unexpected modest negative correlation was found between nadir CD4+ T cell counts and spike-specific CD8+ T cell responses after both the second (Spearman’s ρ=-0.29, p=0.04) and third (Spearman’s ρ=-0.28, p=0.05) dose. We hypothesize that the post-third-dose results are because PLWH with a history of immune dysfunction were eligible for a higher (100 mcg) third dose of mRNA-1273, though this would not explain our post-second-dose results. Regardless, there is no indication that a low nadir CD4+ count impaired T cell responses to COVID-19 vaccination in our cohort.

### Breakthrough infection further boosts CD4+ and CD8+ T cell responses

In British Columbia, the initial waves of Omicron BA.1 and BA.2 occurred after the mass administration of third vaccine doses in the province. Of the 50 PLWH studied, 21 (42%) PLWH experienced SARS-CoV-2 breakthrough infections between 1 and 6 months following their third dose. Post-breakthrough plasma and PBMC specimens were available from nearly all PLWH, so we further examined humoral (neutralizing antibody) and cellular immune responses 6 months after the third dose, and again one month after receipt of a fourth vaccine dose, in the PLWH cohort (n=50) (**Figure 3**). As we reported previously [12, 24], plasma neutralizing antibody activity declined significantly among SARS-CoV-2-naïve individuals after the third vaccine dose, but in those who experienced breakthrough infection, neutralization activity increased to above peak levels induced by vaccination. For example, at 6 months post-third vaccine dose, the median virus neutralization titer (reported as a reciprocal dilution) was 640 (IQR 320-2560) in PLWH who experienced a breakthrough infection, double the observed value in this group after three vaccine doses alone (paired test, p=0.002; **Figure 3A**). This neutralization titer was also eight times higher than the median titer of 80 (IQR 20-160) observed at 6 months post-third vaccine dose in PLWH who remained SARS-CoV-2 naïve (p<0.0001; **Figure 3A**). The observation that neutralization titers increase significantly following breakthrough infection confirms the ongoing presence of spike-specific memory B cells despite diminished neutralizing activity in blood.

**Figure 3.**
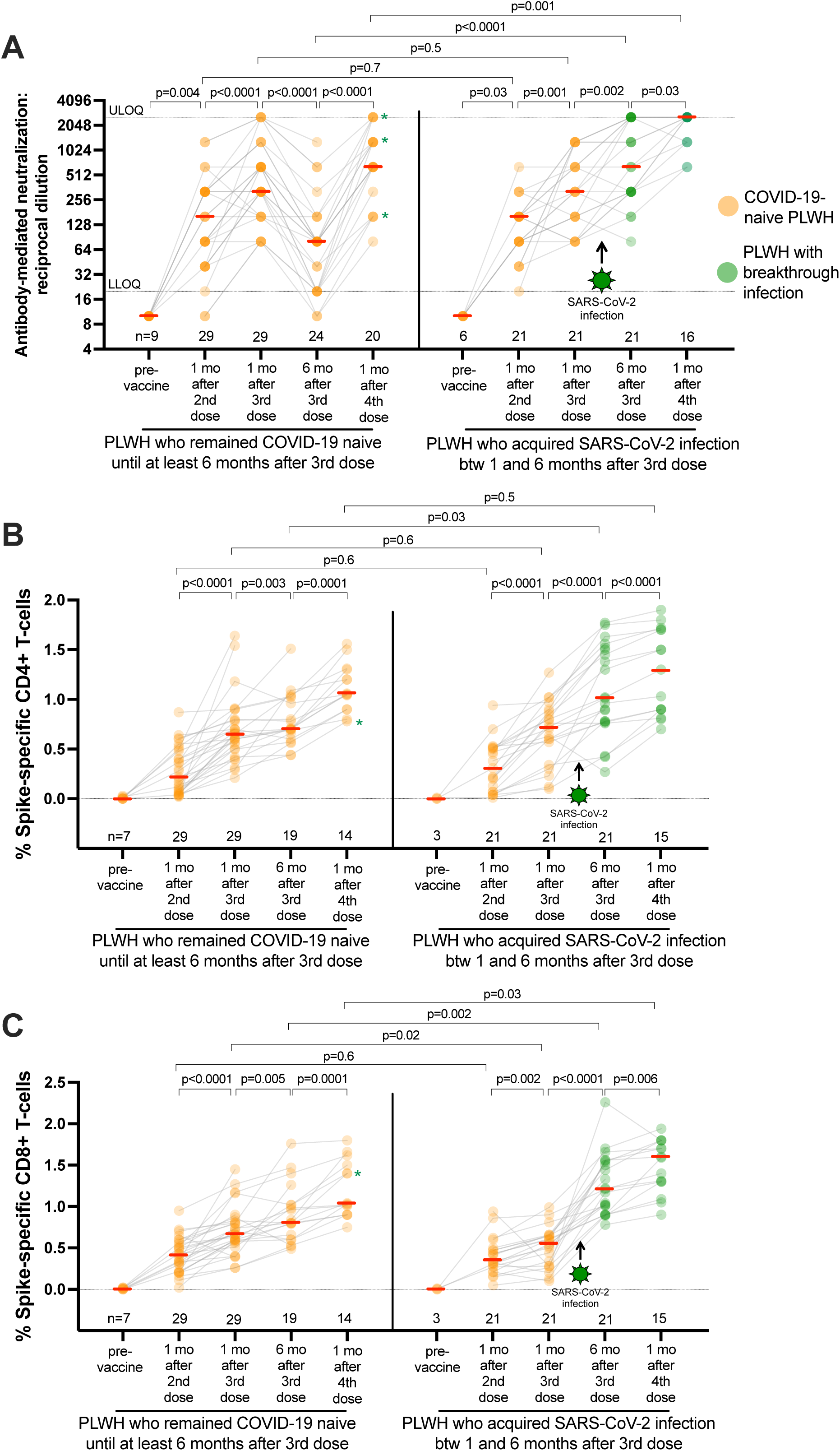
Longitudinal SARS-CoV-2 spike-specific immune responses in PLWH who remained naïve or who experienced their first SARS-CoV-2 infection after COVID-19 vaccination. *Panel A:* SARS-CoV-2 neutralizing antibody titers (reciprocal dilution) in people living with HIV (PLWH) after two, three and four doses of COVID-19 vaccine. Data from the same individual are connected by grey lines. SARS-CoV-2-naïve participants are displayed in orange. SARS-CoV-2 breakthrough participants after the third dose are displayed in green (starting at the point of seroconversion). Note that many antibody titer values overlap, resulting in darker colored symbols. Asterisks denote naive participants who experienced SARS-CoV-2 breakthrough between six months post-dose three and one month post-dose four. Red bars indicate median. The Mann-Whitney U-test was used for between-group comparisons and the Wilcoxon matched pairs test was used for longitudinal paired comparisons. P-values are not corrected for multiple comparisons. *Panel B*: Same as panel A, but for spike-specific CD4+ T cell responses. *Panel C:* Same as panels A and B, but for spike-specific CD8+ T cell responses. The Mann-Whitney U-test was used for between-group comparisons and the Wilcoxon matched pairs test was used for longitudinal paired comparisons. P-values are not corrected for multiple comparisons.

In contrast to neutralizing antibodies, spike-specific CD4+ and CD8+ T cell frequencies did not wane in SARS-CoV-2 naïve PLWH following vaccination. In fact, the percentage of both spike-specific CD4+ and CD8+ T cells increased slightly between 1 and 6 months after the third vaccine dose (paired p=0.003 and p=0.005 respectively, **Figures 3B and 3C**), consistent with the continued presence of vaccine-induced T cells even in the absence of antigen. Like humoral responses however, breakthrough infection boosted T cell responses to levels higher than those observed in the same participants after vaccination alone (both paired tests p≤0.0001; **Figures 3B and 3C**). Moreover, the median percentage of spike-specific CD4+ T cells at 6 months post-third vaccine dose in breakthrough cases was a 1.01% (IQR 0.78-1.51), compared to 0.70% (IQR 0.60-1.02) in SARS-CoV-2-naïve participants at this same time point (p=0.03; **Figure 3B**), while the median percentage of spike-specific CD8+ T cells in breakthrough cases was 1.20% (IQR 0.96-1.55), compared to 0.80% (IQR 0.60-1.02) in SARS-CoV-2 naïve participants at this time point (p=0.002; **Figure 3C**).

A fourth vaccine dose significantly increased neutralizing antibody responses and spike-specific T cell responses in SARS-CoV-2 naïve PLWH (all paired tests p≤0.0001; **Figures 3A, 3B, 3C**), illustrating the immune benefit of a fourth vaccine dose in the absence of SARS-CoV-2 infection. A fourth dose also significantly increased humoral and cellular immune responses in PLWH who experienced prior breakthrough infection (all paired tests p<0.03; **Figures 3A, 3B, 3C**). Of note, post-fourth dose neutralizing antibody titers and CD8+ T cell responses in PLWH who experienced breakthrough infection were significantly higher than among SARS-CoV-2 naïve PLWH at this same time (p=0.001 and p=0.03 respectively; **Figures 3A and 3C**), consistent with prior reports of superior “hybrid” immunity generated by a combination of vaccination and infection [31, 32].

### Correlates of protection against breakthrough infection

Given the substantial proportion of participants who experienced their first SARS-CoV-2 breakthrough infection between one and six months following three-dose vaccination (42% of PLWH; 44% of controls), we explored our data for potential correlates of protection. As HIV status was not associated with breakthrough infection (p=0.86), PLWH and control groups were combined for analysis, yielding 78 participants who remained SARS-CoV-2 naïve and 59 participants who experienced SARS-CoV-2 infection during the follow-up period. Univariable analyses revealed older age to be significantly associated with protection against breakthrough infection: participants remaining SARS-CoV-2 naïve were a median 60 [IQR 42-71] years old while those who experienced breakthrough infection were a median 43 [IQR 34-60] years old (combined cohort p=0.001; **Figure 4A**). This observation was largely driven by the control group, which included 29 adults age 70 years or over (compared to 3 in the PLWH group). The percentage of spike-specific CD4+ T cells one month after the third vaccine dose was not associated with breakthrough infection (p=0.72 for the combined cohort; **Figure 4B**), but a higher percentage of spike-specific CD8+ T cells at this time point was associated with remaining SARS-CoV-2 naïve (p=0.046 for the combined cohort) (**Figure 4C**). This observation was driven by differences in CD8+ T cell responses among PLWH (p=0.02) and suggests that cytotoxic T cells specific for SARS-CoV-2 may contribute to protection. Somewhat surprisingly, we found no association between neutralizing antibody responses induced by three vaccine doses and breakthrough infections (p=0.33 for the combined cohort) (**Figure 4D**), likely because vaccine-induced antibodies against the ancestral (Wuhan) spike had limited activity against the Omicron strains that caused the majority of breakthrough infections, and because these infections occurred a median of 3.7 months after booster vaccination, when circulating antibody titers had declined substantially. In a multivariable logistic regression analysis that incorporated participant sociodemographic, vaccine and immune response variables, older age was the only significant predictor of protection against SARS-CoV-2 breakthrough infection (**Table S3**). Though at first surprising, this observation is consistent with the very rapid rise in SARS-CoV-2 anti-N seroprevalence in Canada among younger individuals following the emergence of Omicron, and likely reflects the impact of social/behavioural factors rather than biological factors. For example, by mid-June 2022, anti-N seroprevalence in Canadian adults aged < 25 years was 57%, in those aged 25–39 years it was 51%, in those aged 40–59 years it was 40%, and in those 60 years or older it was 25% [33]. Similar age-specific trends were observed in Finland [34] and the USA [35], suggesting that many older adults continued to benefit from efforts to limit exposure to COVID-19 in the community, even after the arrival of Omicron.

**Figure 4.**
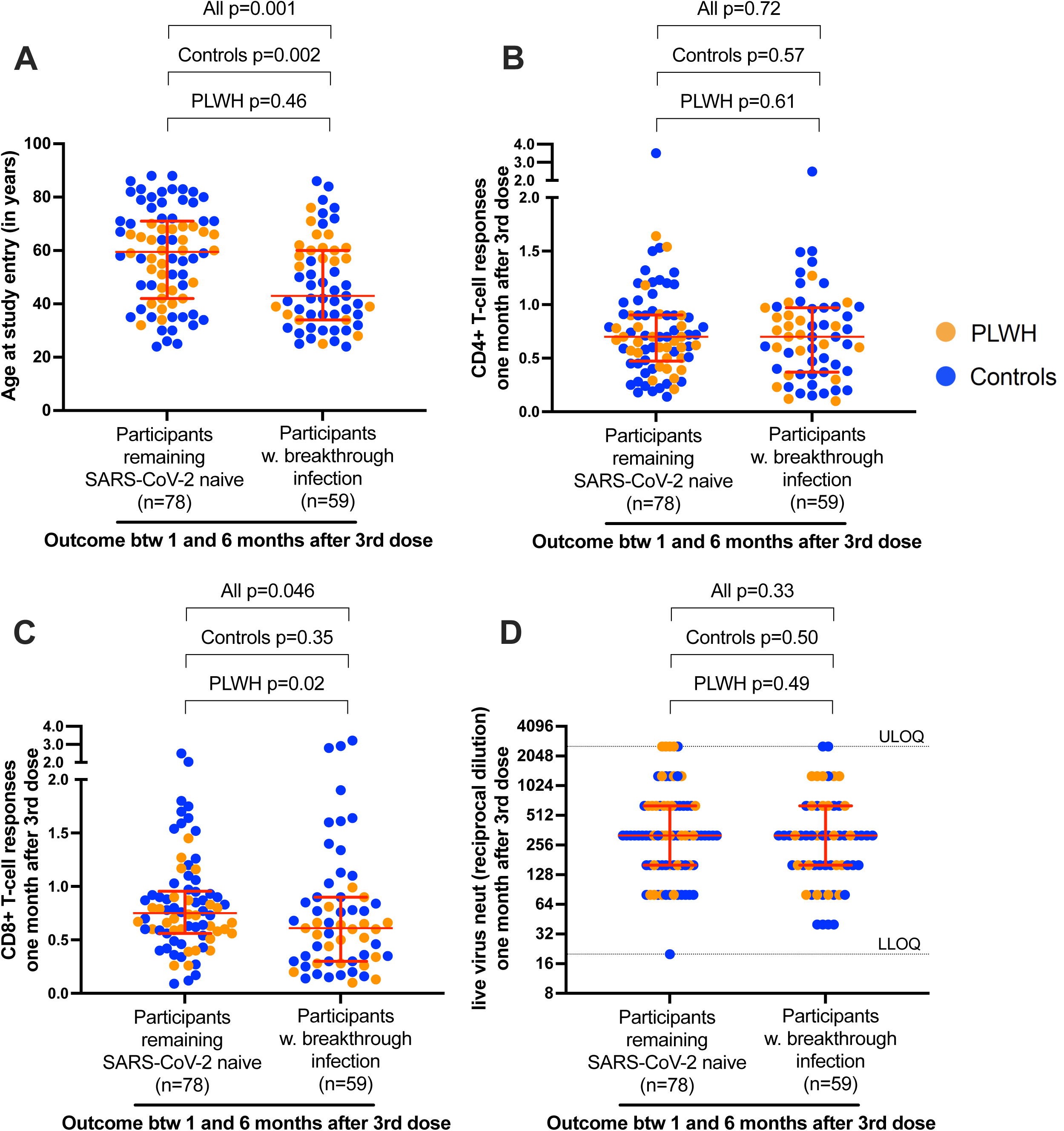
Analysis of correlates of protection against SARS-CoV-2 breakthrough infection after COVID-19 vaccination. *Panel A:* Participant ages are shown at study entry, stratified according to those who remained SARS-CoV-2 naive (n=78) and those who experienced SARS-CoV-2 breakthrough infection between one and six months after receiving a third vaccine dose (n=59). PLWH are in orange; controls are in blue. *Panel B:* Same as panel A, except the frequencies of spike-specific CD4+ T cells one month after three vaccine doses are shown. *Panel C:* Same as panel A and B, except the frequencies of CD8+ T cells one month after three vaccine doses are shown. *Panel D:* Same as other panels, except SARS-CoV-2 neutralization activities (reported as reciprocal dilution titers) one month after three vaccine doses are shown. For all panels, the Mann-Whitney U-test was used to compare groups, using data from the full cohort (all; n=137), from controls only (n=87), or from PLWH only (n=50).

## DISCUSSION

We observed that the frequencies of SARS-CoV-2 spike-specific CD4+ and CD8+ T cells were not significantly different between PLWH and controls after two doses of COVID-19 vaccine. Both CD4+ and CD8+ T cell responses were enhanced following a third vaccine dose, but the frequency of spike-specific CD8+ T cells was modestly lower in PLWH compared to controls. PLWH generated robust spike-specific T cell responses following post-vaccine breakthrough infection, reaching levels that were equivalent to those achieved after a fourth vaccine dose in SARS-CoV-2 naïve PLWH. Of note, receiving a fourth vaccine dose after having experienced a breakthrough SARS-CoV-2 infection enhanced immune responses even further, with neutralizing antibody responses and CD8+ T cell responses reaching levels significantly higher than those observed in SARS-CoV-2 naïve individuals after four doses. This finding is consistent with previous reports describing the superiority of “hybrid” immunity generated through a combination of vaccination and infection [31, 32].

Our results are broadly consistent with other recent studies showing that vaccine-induced spike-specific T cell responses are equivalent in PLWH and controls without HIV, particularly among PLWH receiving antiretroviral therapy [9, 17–20, 23]. We found no association between CD4+ T cell responses and neutralizing antibodies after either two or three vaccine doses, suggesting that sufficient T cell help was provided to stimulate spike-specific B cell responses despite a wide range of antigen-specific CD4+ T cell frequencies induced by vaccination. While our results do not rule out potential dysfunction in vaccine-induced T cells among PLWH, our data suggest that the impact of T cell impairment is likely to be modest after the second and third vaccine dose.

This study has some limitations. All PLWH were receiving suppressive antiretroviral therapy and had generally preserved CD4+ T cell counts, so our observations may not be generalizable to PLWH who have low CD4+ T cell counts or untreated HIV. Since this was an observational study, PLWH and control participants were not matched at enrolment. Though we included known variables in multivariable regression analyses, residual effects of variables that differed between groups (e.g. sex, vaccine regimen type) or other unknown factors could influence our results. We also lacked information on the concentration of mRNA-1273 third doses (100 mcg versus 50 mcg), and therefore we could not investigate the impact of this on immune responses.

While the activation induced marker assay provides a sensitive method to quantify spike-specific CD4+ and CD8+ T cells, we did not analyze T cell subsets such as follicular helper cells, nor T cell functions such as cytokine production or proliferation, that could reveal differences in the effector activity of vaccine-or infection-induced responses. Finally, since T cell responses were measured using peptide pools spanning only the ancestral spike protein, we cannot comment on the relative dominance of individual T cell epitopes nor on potential changes in the distribution of T cell responses following repeated vaccination or breakthrough infection.

## CONCLUSIONS

In summary, our study indicates that PLWH who are receiving suppressive antiretroviral therapy mount strong CD4+ and CD8+ T cell responses to COVID-19 vaccines that are comparable to those found in individuals without HIV. Repeated exposure to SARS-CoV-2 spike antigen, either through vaccination or infection, further enhanced T cell responses in PLWH. PLWH status was not associated with increased risk of post-vaccine breakthrough infection in our cohort, but older age was found to be a significant predictor of protection against infection, consistent with lower SARS-CoV-2 anti-N seropositivity rates among older adults in Canada following the emergence of Omicron [33]. These results further underscore the benefits of COVID-19 vaccination in PLWH.

## Data Availability

The datasets analysed in this study are available from the corresponding author upon reasonable request. Data have also been deposited into the COVID-19 Immunology Task Force (CITF) data bank, available online at: https://portal.citf.mcgill.ca.

## DECLARATIONS

### Consent for publication

Not applicable.

### Availability of data and materials

The datasets analysed in this study are available from the corresponding author upon reasonable request. Data have also been deposited into the COVID-19 Immunology Task Force (CITF) data bank (https://portal.citf.mcgill.ca/).

### Authors’ contributions

SD, ZLB and MAB led the study. YS and HRL coordinated the study. SD and RK quantified T cell responses by flow cytometry. SD, RK, RW, FM, YS, SS, NM-G, MCD, FHO and EB processed specimens and collected data. PKC performed viral neutralization assays. SD, RK, MAB and ZLB analyzed data. RL performed statistical analyses. MLD and MGR supervised SARS-CoV-2 serum antibody quantification. MH, CTC, MH, MH, MGR and JGSM contributed to cohort establishment. ZLB and MAB drafted the manuscript. All authors contributed to manuscript review and editing.

## Acknowledgements

We thank the leadership and staff of Providence Health Care, St. Paul’s Hospital, the BC Centre for Excellence in HIV/AIDS, the Hope to Health Research and Innovation Centre, and Simon Fraser University for supporting this project. In particular, we thank Landon Young and Bruce Ganase for phlebotomy assistance; Laura Burns for technical assistance; Dr Junine Toy, Dr. Chanson Brumme and Paul Sereda for database assistance; and Drs. Silvia Guillemi, Rolando Barrios, Victor Leung, Janet Simons, Daniel Holmes, Curtis Cooper and Aslam Anis for clinical support and guidance. Finally, we thank the participants, without whom this study would not have been possible.

---

**TABLE S1.** Multivariable zero-inflated beta regressions exploring the relationship between sociodemographic, health and vaccine-related variables on Spike-specific CD4+ T cell responses following two and three COVID-19 vaccine doses.

| Variable | Outcome measures |  |  |  |  |  |
| --- | --- | --- | --- | --- | --- | --- |
|  | % Spike-responsive CD4+ T cells<br>after two vaccine doses |  |  | % Spike-responsive CD4+ T cells<br>after three vaccine doses |  |  |
|  | Estimate | Standard<br>error | P value | Estimate | Standard<br>error | P value |
| PLWH group | -0.29 | 0.17 | 0.08 | -0.06 | 0.10 | 0.58 |
| Age (per year) | 0.001 | 0.004 | 0.83 | -0.004 | 0.002 | 0.08 |
| Male sex | 0.13 | 0.14 | 0.38 | 0.24 | 0.084 | **0.0044** |
| White ethnicity | -0.19 | 0.14 | 0.16 | -0.045 | 0.076 | 0.55 |
| Health conditions (per condition)<sup>a</sup> | -0.03 | 0.08 | 0.72 | 0.11 | 0.047 | **0.022** |
| ChAdOx1-containing initial vaccine regimen<sup>b</sup> | 0.37 | 0.21 | 0.09 | 0.14 | 0.12 | 0.27 |
| Days between 1st and 2nd vaccine dose (per day) | 0.005 | 0.003 | 0.12 | 0.002 | 0.002 | 0.30 |
| mRNA-1273 as third vaccine dose<sup>c</sup> | N.A. | N.A. | N.A. | 0.17 | 0.077 | **0.033** |
| Days between 2nd and 3rd vaccine dose (per day) | N.A. | N.A. | N.A. | 0.001 | 0.001 | 0.47 |
| % Spike specific T cells after two doses (per 1% increment) | N.A. | N.A. | N.A. | 0.71 | 0.058 | **<2x10<sup>-16</sup>** |

<sup>a</sup>Continuous variable, defined as: chronic blood disorder, cancer, hypertension, diabetes, asthma, obesity (body mass index $\geq 30$ ), chronic diseases of lung, liver, kidney or heart
<sup>b</sup>Binary variable, defined as two doses of ChAdOx1, or a heterologous regimen consisting of one dose each of ChAdOx1 and an mRNA vaccine (mRNA-1273 or BNT162b2) (vs. a reference group defined as two doses of mRNA vaccine)
<sup>c</sup>Binary variable, defined as a third dose of mRNA-1273 (vs. a reference group defined as a third dose of BNT162b2)

**TABLE S2.** Multivariable zero-inflated beta regressions exploring the relationship between sociodemographic, health and vaccine-related variables on Spike-specific CD8+ T cell responses following two and three COVID-19 vaccine doses.

| Variable | Outcome measures |  |  |  |  |  |
| --- | --- | --- | --- | --- | --- | --- |
|  | % Spike-responsive CD8+ T cells after two vaccine doses |  |  | % Spike-responsive CD8+ T cells after three vaccine doses |  |  |
|  | Estimate | Standard error | P value | Estimate | Standard error | P value |
| PLWH group | -0.0002 | 0.16 | 0.99 | -0.24 | 0.12 | **0.045** |
| Age (per year) | -0.001 | 0.004 | 0.87 | 0.001 | 0.004 | 0.87 |
| Male sex | 0.18 | 0.14 | 0.20 | 0.11 | 0.10 | 0.24 |
| White ethnicity | 0.14 | 0.13 | 0.30 | -0.01 | 0.08 | 0.90 |
| Health conditions (per condition)<sup>a</sup> | -0.02 | 0.08 | 0.75 | -0.05 | 0.06 | 0.37 |
| ChAdOx1-containing initial vaccine regimen<sup>b</sup> | 0.39 | 0.20 | **0.049** | 0.22 | 0.14 | 0.12 |
| Days between 1st and 2nd vaccine dose (per day) | 0.002 | 0.003 | 0.54 | 0.00 | 0.00 | 0.16 |
| mRNA-1273 as third vaccine dose<sup>c</sup> | N.A. | N.A. | N.A. | 0.02 | 0.11 | 0.86 |
| Days between 2nd and 3rd vaccine dose (per day) | N.A. | N.A. | N.A. | -0.001 | 0.002 | 0.43 |
| % Spike specific T cells after two doses (per 1% increment) | N.A. | N.A. | N.A. | 0.79 | 0.07 | **<2e-16** |

<sup>a</sup>Continuous variable, defined as: chronic blood disorder, cancer, hypertension, diabetes, asthma, obesity (body mass index $\geq 30$ ), chronic diseases of lung, liver, kidney or heart
<sup>b</sup>Binary variable, defined as two doses of ChAdOx1, or a heterologous regimen consisting of one dose each of ChAdOx1 and an mRNA vaccine (mRNA-1273 or BNT162b2) (vs. a reference group defined as two doses of mRNA vaccine)
<sup>c</sup>Binary variable, defined as a third dose of mRNA-1273 (vs. a reference group defined as a third dose of BNT162b2)

**TABLE S3.**
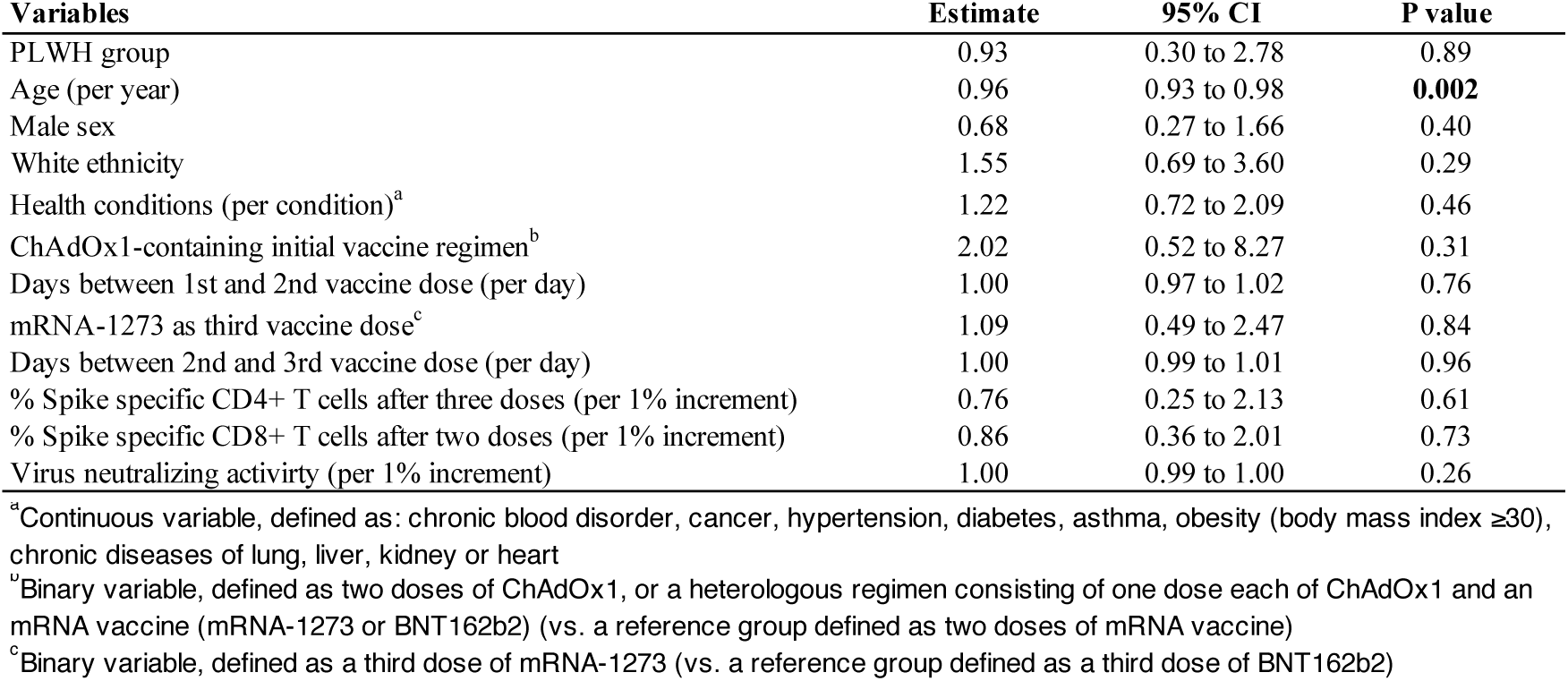
Multivariable logistic regression looking at factors associated with risk of first breakthrough infection between 1 and 6 months following receipt of the 3rd COVID-19 vaccine dose.

**Figure S1.**
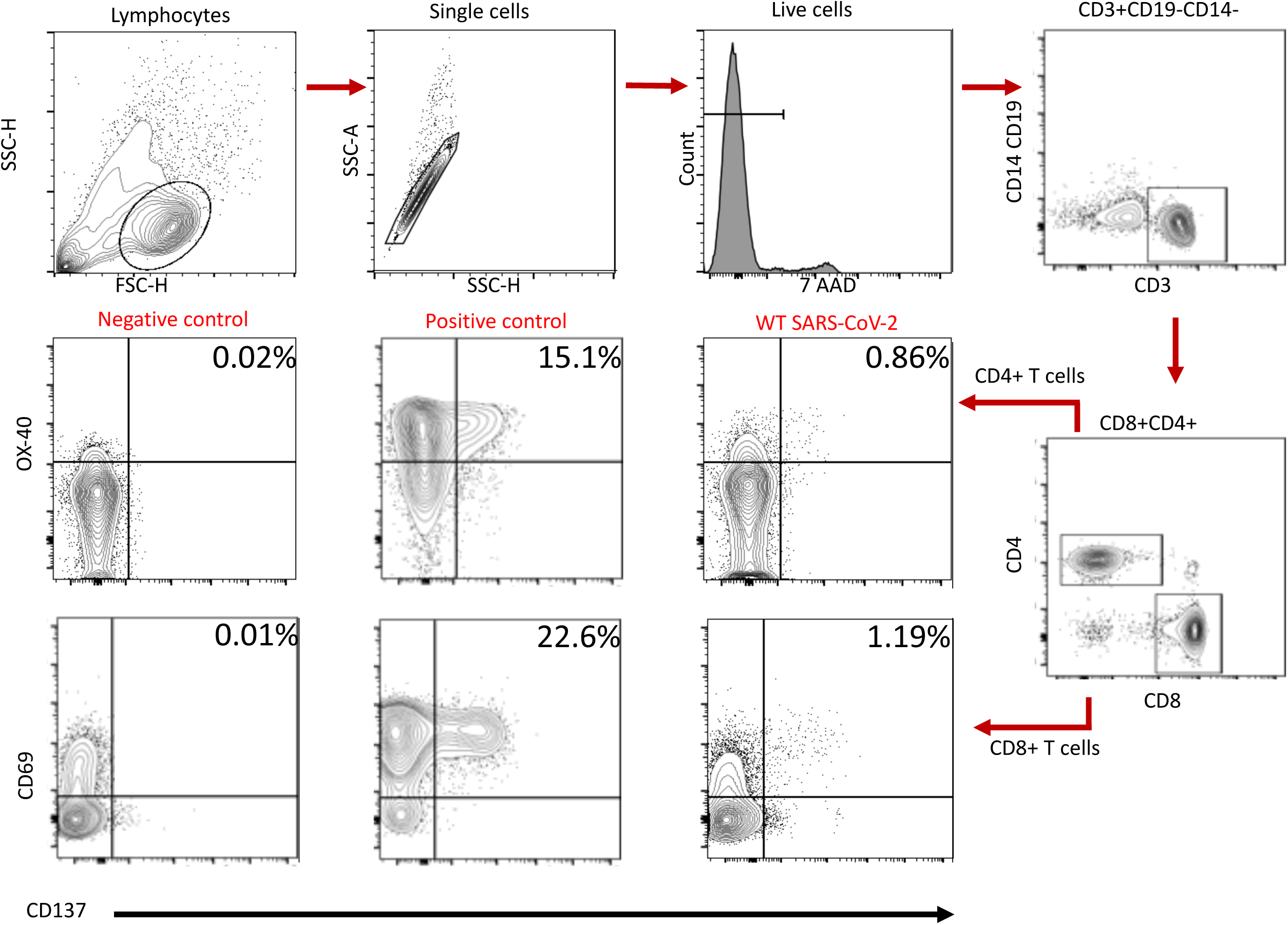
Gating strategy used to quantify vaccine-induced spike-specific T cell responses.

**Figure S2.**
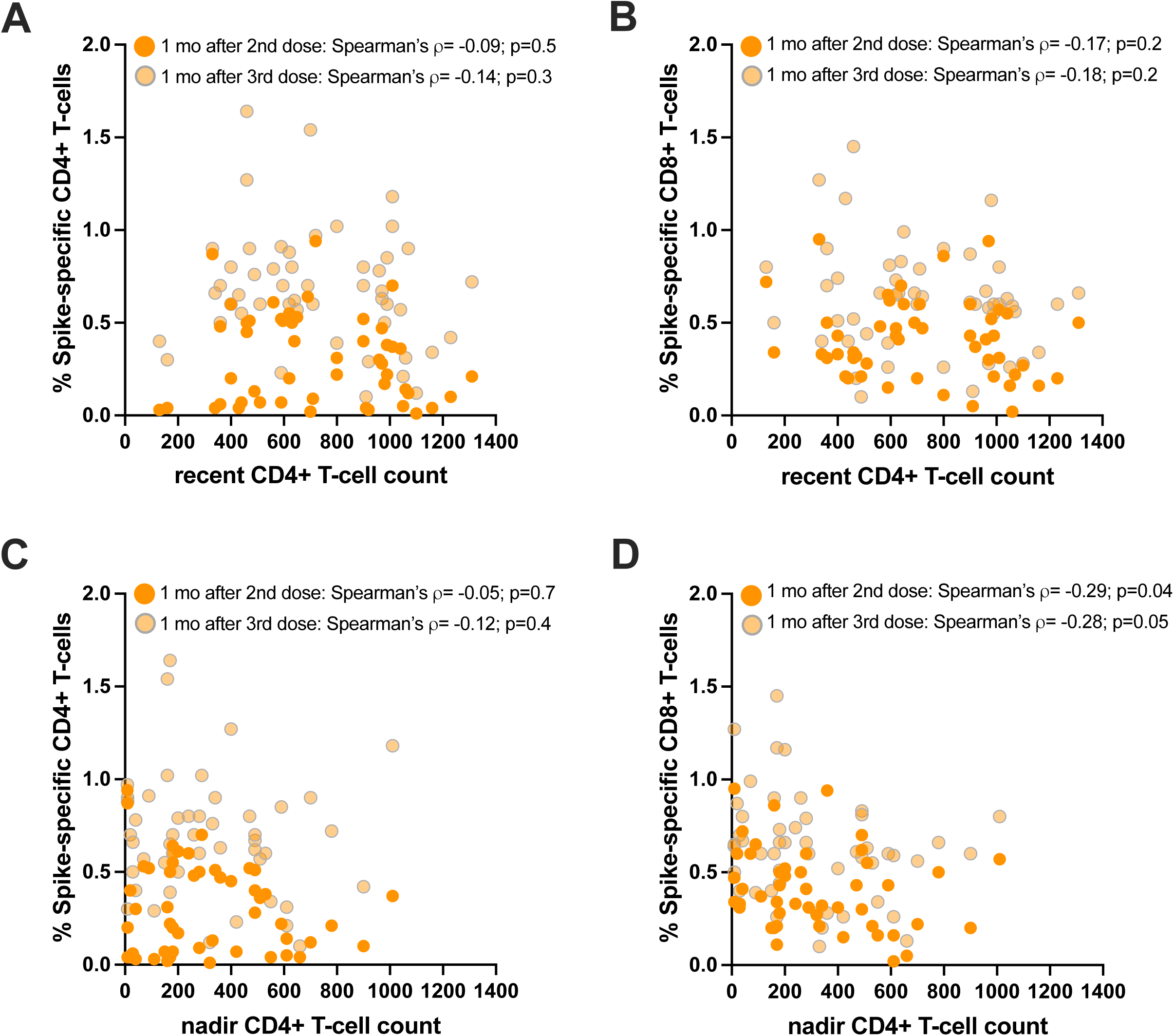
Correlations between vaccine-induced spike-specific T cell responses and recent or nadir.

